# A Preliminary Study of Using Computer Vision to Quantify Trunk Recruitment during Bimanual Play and the Effect of Upper Extremity Interventions in Unilateral Spastic Cerebral Palsy

**DOI:** 10.64898/2026.01.11.26343639

**Authors:** Dalina Delfing, Shivakeshavan Ratnadurai-Giridharan, Karen Chin, Kathleen M. Friel, Andrew M. Gordon

## Abstract

**Background:** Children with unilateral spastic cerebral palsy (USCP) often rely on trunk compensation due to impaired upper limb control, but current clinical tools do not directly capture trunk involvement. Marker-based systems are challenging to use with children, while computer vision methods like OpenPose offer a promising, scalable alternative for kinematic analysis but need to be validated.

**Purpose:** We validated OpenPose for quantifying trunk recruitment during bimanual play in children with USCP and examined how the interventions Constraint-Induced Movement Therapy (CIMT) and Hand-Arm Bimanual Intensive Therapy (HABIT) influence trunk use.

**Methods:** We analyzed videos of children with USCP who underwent CIMT or HABIT. OpenPose was used to extract trunk displacement angle (TDA) and trunk rotation angle (TRA), which were compared to hand function scores. OpenPose was validated against a 3D motion analysis system in typically developing adults. Reach-phase kinematic variables were also assessed.

**Results:** OpenPose showed high validity for TDA and lower validity for multi-planar TRA. TDA and TRA did not correlate with baseline hand function. HABIT reduced TDA, while CIMT slightly increased it. No significant changes were found in velocity, movement time, or variability.

**Conclusions:** OpenPose is a viable tool for capturing gross trunk motion. Trunk recruitment patterns differed by intervention, supporting the need for personalized approaches.

## 5.1 Introduction

Human movements are complex and adaptable, allowing us to adjust to various environmental conditions and movement challenges (Jaspers et al., 2011; Klingels et al., 2012; Steenbergen & Van Der Kamp, 2004). A specific condition that often requires adapting is cerebral palsy (CP), a non-progressive disorder causing motor impairments due to brain damage or abnormal development before, during, or shortly after birth (Rosenbaum et al., 2007). Unilateral spastic cerebral palsy (USCP), also known as hemiplegia, is the most common subtype of CP and is characterized by movement impairments predominantly affecting one side of the body, with the upper body being more severely impacted (Sadowska et al., 2020). The more affected upper limb exhibits reduced range of motion in the elbow and shoulder, muscle weakness, spasticity causing muscle shortening and decreased elbow angle, more pronounced distal than proximal motor impairments, and compromised motor control and coordination (Jaspers et al., 2011; Klingels et al., 2012; Steenbergen & Van Der Kamp, 2004). Additionally, Santamaria et al., (2020) demonstrated that impaired reaching in children with moderate to severe CP is often associated with inadequate trunk stabilization.

These impairments greatly affect daily activities such as prehension. Proximal muscles of the trunk, shoulder, and elbow guide hand transport and positioning while distal muscles control grasping (Coluccini et al., 2007; Jeannerod, 1988; Kuhtz-Buschbeck et al., 1998). Typically, during early motor development, proximal and distal joints form a synergy to reduce the degrees of freedom the brain must control. With practice, this synergy becomes increasingly flexible and dynamic, engaging the trunk muscles based on target distance (Cirstea & Levin, 2000). However, in individuals with USCP, trunk movements appear to be a primary component of their synergy rather than merely a compensatory response. This is supported by studies showing that the trunk is recruited even when its involvement is biomechanically unnecessary, suggesting that trunk use reflects a reorganized movement pattern. This reorganization may be shaped by early motor impairments and by a tendency to reduce the effort required to engage the full capacity of distal joints (Cirstea & Levin, 2000; Kreulen et al., 2007; Meulenbroek, 2006; Roby-Brami et al., 1997). Additionally, unlike healthy individuals who dynamically utilize all three trunk dimensions, namely forward bending, inclination angling, and rotation, USCP patients primarily adjust forward bending while inclination and rotation remain consistent across tasks (Kreulen et al., 2007; Meulenbroek, 2006). Individuals with more severe distal joint deficits tend to rely more on trunk compensation during reaching tasks. Trunk involvement helps position the upper limb and compensates for impairments such as difficulty with forearm supination. By recruiting the trunk, the shoulder and elbow require fewer degrees of freedom. This reduces reliance on distal joints and brings the hand closer to the target, allowing for more stable interaction with objects. (Cirstea & Levin, 2000; Darji & Diwan, 2022; Levin et al., 2016; Yildiz et al., 2018).

Constraint-Induced Movement Therapy (CIMT) and Hand-Arm Bimanual Intensive Training (HABIT) have been used successfully to improve upper limb motor function in USCP (Gordon et al., 2011; Hung et al., 2018; Novak & Honan, 2019; Sakzewski et al., 2010). CIMT involves restraining the less affected limb, encouraging practice with the more affected limb, while HABIT targets coordinated bilateral limb use (Friel et al., 2021). Interestingly, research into intervention effects on trunk involvement has found ambiguous results. In stroke, a condition with very similar motor impairments, research indicates increased trunk use post-CIMT (Kitago et al., 2013; Massie et al., 2009). Conversely, pediatric USCP studies show CIMT either reduces or does not affect trunk involvement (Hung et al., 2020; Shih et al., 2023). HABIT diminished shoulder displacement in USCP children (Hung et al., 2020). These discrepancies exist because of limited research, diverse methodologies, and inconsistent measurement approaches, highlighting the need for more research to clarify the relationship between rehabilitation interventions and trunk recruitment in individuals with USCP.

Current clinical scales, such as the Assisting Hand Assessment (AHA), the gold standard for bimanual function in USCP, do not specifically capture trunk contribution. While 3D motion analysis (3DMA) is insightful, it requires expensive specialized equipment, technical expertise, and laboratory space, limiting its accessibility (Mathis et al., 2020). Deep learning algorithms offer cost-effective solutions because they can analyze large datasets directly and are marker-less, reducing costs, discomfort, and extending research beyond the lab setting (Cronin, 2021; Mathis et al., 2020). OpenPose, a deep learning algorithm, has been shown to accurately predict clinically relevant metrics such as walking speed, cadence, and knee flexion angle in CP research (Kidziński et al., 2020; Stenum et al., 2021).

### 5.1.1 Study Aims

In the present study we conducted a retrospective analysis of AHA hand function videos to determine the relationship between trunk displacement (TDA) and trunk rotation angle (TRA) with upper extremity function using OpenPose. To ensure the validity of this approach, we first validated OpenPose for our intended application by comparing its outputs against a gold-standard 3D motion analysis system (3DMA) in typically developing adults performing reaching tasks. We hypothesized that TDA and TRA would correlate negatively with baseline hand function tests, indicating greater trunk compensation in children with lower upper extremity function. Additionally, we explored velocity, movement time, and variability during the reach, grasp, and transport phases to assess their contribution to hand function and trunk involvement. We also examined how CIMT and HABIT impacted trunk involvement. We hypothesized that CIMT would increase TDA and TRA, while HABIT would decrease it, reflecting intervention-specific movement adaptations. The results of this study may help to tailor interventions to individual needs, increasing participation in daily activities.

## 5.2 Method

### 5.2.1 Trunk Recruitment Study

#### Participants

Retrospective analyses included 50 children aged 6-17 years with USCP (29 males; Mean age=9.28 yrs, SD=3.08). Participants from previous clinical trial underwent CIMT/HABIT intervention, approved under IRB Protocol #14-140. For more information on study recruitment, see (Friel et al., 2021), https://doi.org/10.3389/fneur.2021.660780.

#### Interventions

Participants attended a three-week day camp at Teachers College, Columbia University, randomly assigned to CIMT or HABIT for six hours of therapy per day, five days per week. Both interventions utilized whole-task practice involving successive, coordinated movements within activities (games, arts and crafts, goal training) lasting 15-20 minutes each. Part-task (shaping) activities increased intensity by maximizing repetitions within at least five 30-second intervals, progressing when the child achieved ≥80% success.

In the CIMT intervention, a cotton sling was attached to the child’s trunk, enclosing the less affected arm to prevent its usage. Task choice consisted of a list of fine motor and manipulative gross motor activities that induced the movement behaviors targeted, which were predefined by interventionists and supervisors. These activities were unimanual functional and play activities that were age-appropriate and targeted specific hand impairments. Progression of task difficulty was translated into greater speed, accuracy, and movement repetitions.

In the HABIT intervention, age-appropriate fine and gross motor activities requiring the use of both hands were selected. The activities were chosen by the interventionist, with the goal of increasing complexity from passive assist to manipulator of the more affected arm. Task demands were modified to ensure success.

#### Hand Function Measures

Unimanual dexterity was measured by the BBT and JTHFT on both the more affected and less affected hand. The BBT measures the number of small wooden blocks moved from one side of a box to the other in 60 seconds (Mathiowetz et al., 1985). The JTHFT quantifies time (in seconds) taken to complete six unimanual fine motor tasks, including flipping index cards, small object manipulation, and manipulating cans. It takes approximately 20 minutes to complete. The AHA (version 5.0) quantifies effectiveness of the child’s more affected hand during bimanual activities in a play-like session. Two interchangeable versions (fortress and alien themes) consist of 14 play activities scored on 20 items, including amount of use and grasp type. Videos are reviewed and scored by a certified evaluator blinded to group allocation. All three tests have been proven to be reliable and valid (Araneda et al., 2019; Krumlinde-Sundholm et al., 2007).

#### Set Up and Procedure of AHA Testing

During AHA pre and post assessments, participants were recorded by a 45-degree angled frontal camera, capturing the child’s hands and face and the activity, which each video being approximately 15-25 minutes long. Children were seated at arm’s length from the edge of a table with the instructor sitting across from them.

### 5.2.2 Validation Study

#### Participants

Nine typically developing adults age between 18-60yrs (4 males; Age: M=30.28 yrs, SD=5.80 yrs; range: 22-41 yrs) with no known neurological or motor impairments participated after informed consent was obtained and the study was approved by Teachers College IRB (Protocol #24-304).

#### Experimental Tasks

Two tasks were used to evaluate motion analysis validity between VICON (Workstation 612, Lake Forest, CA) and OpenPose. A circular chart (75 x 110 cm) replicated the AHA test workspace. For the unimanual task, a small box containing a music machine from the AHA test kit was selected, while for the bimanual task, two wooden cymbals from the AHA kit were utilized.

#### Set Up and Procedure

Participants were videotaped with a 45-degree angled, frontally oriented digital camera (Sony HDR-XR350V, 30 Hz), and simultaneously with a 10-camera 3D motion capture system (VICON Nexus v 2.2.1, Lake Forest, CA). Twenty-two reflective markers were attached to specific anatomical landmarks (forehead, clavicle, C7, T10, lower back, hip, shoulders, biceps, elbows, outer edge of forearms, wrists, index fingers). Dynamic calibration (five reaching movements spaced 30° apart using a T-shaped wand) and static calibration (participants sitting quietly, palms-down) was performed.

Participants sat at a 75 x 110 cm table with hands positioned on the table next to them. During the unimanual task, participants used their preferred hand to pick up an object from the black dot on the circular chart, moved it to a designated location, returned their hand to the start position, reached for the object again, returned it to its original position, and then moved their hand back to start. The bimanual task was similar, except participants simultaneously manipulated two wooden clamps, one in each hand. Both tasks had eight trials using the same task and trial order for each participant (Figure 5.1). Placement locations of the objects were given verbally, and the participants performed the tasks at their own pace. The total time to perform both tasks was approximately three minutes.

**Figure 5.1.**
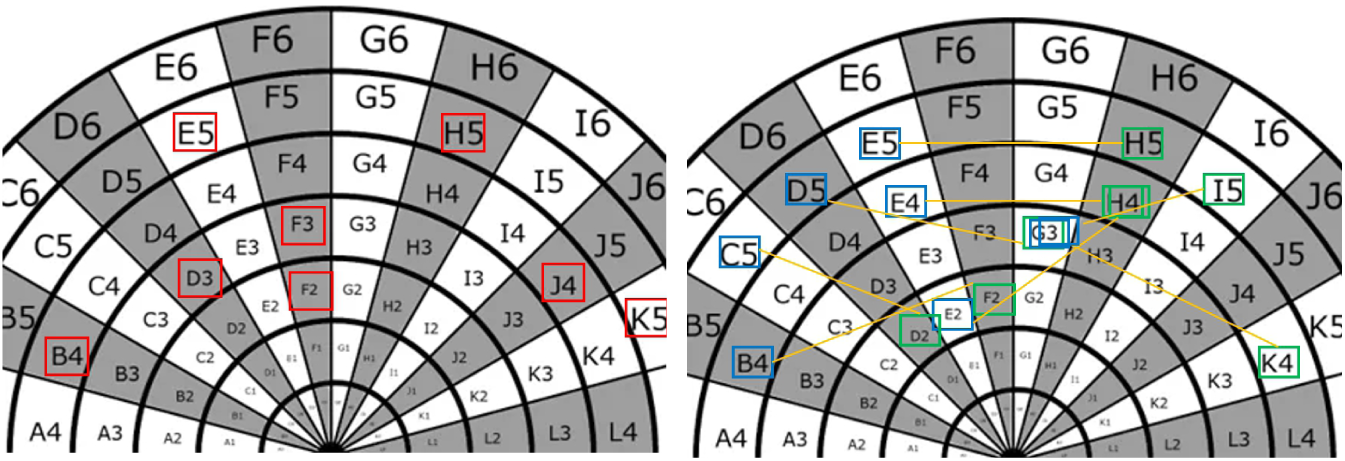
Target Locations for Unimanual and Bimanual Trials. Note. This illustrates the eight targets completed by participants during the unimanual and bimanual trials. On the left, the target locations for the unimanual trials are marked in red. Bimanual trial targets are color-coded on the right side: blue indicates targets for the nonpreferred hand, and green represents targets for the preferred hand. The yellow lines connecting the target locations in the bimanual trials indicate paired targets that participants aimed for simultaneously within the same trial.

### 5.2.3 Data Analysis

#### Pre-processing

##### 3DMA

For the validation part, 3DMA data were manually labeled to ensure accuracy of marker identification. The data were then exported into an Excel CSV file for further processing.

##### 2D Video Recordings

OpenPose, a pre-trained deep learning library for estimating the position of body joints as pixel locations in an image, was used to pre-process 2D videos on high-performance computing nodes, extracting 25 key points per frame. Pre-processing included frame-by-frame detection, iterative optimization, and post-processing refinements. The data were exported to an Excel CSV file.

#### Kinematic Metrics

##### Trunk Recruitment Study

Two trunk metrics were analyzed: 1) Trunk displacement angle (TDA), calculated as the average deviation of the trunk (hip-to-neck vector) from its median position over time; and 2) Trunk rotation angle (TRA), calculated as the median deviation of the shoulder-to-shoulder vector over time. For both metrics, a minimum value of 0 indicated a stiff upper body with no leaning or rotation (Figure 5.2).

**Figure 5.2.**
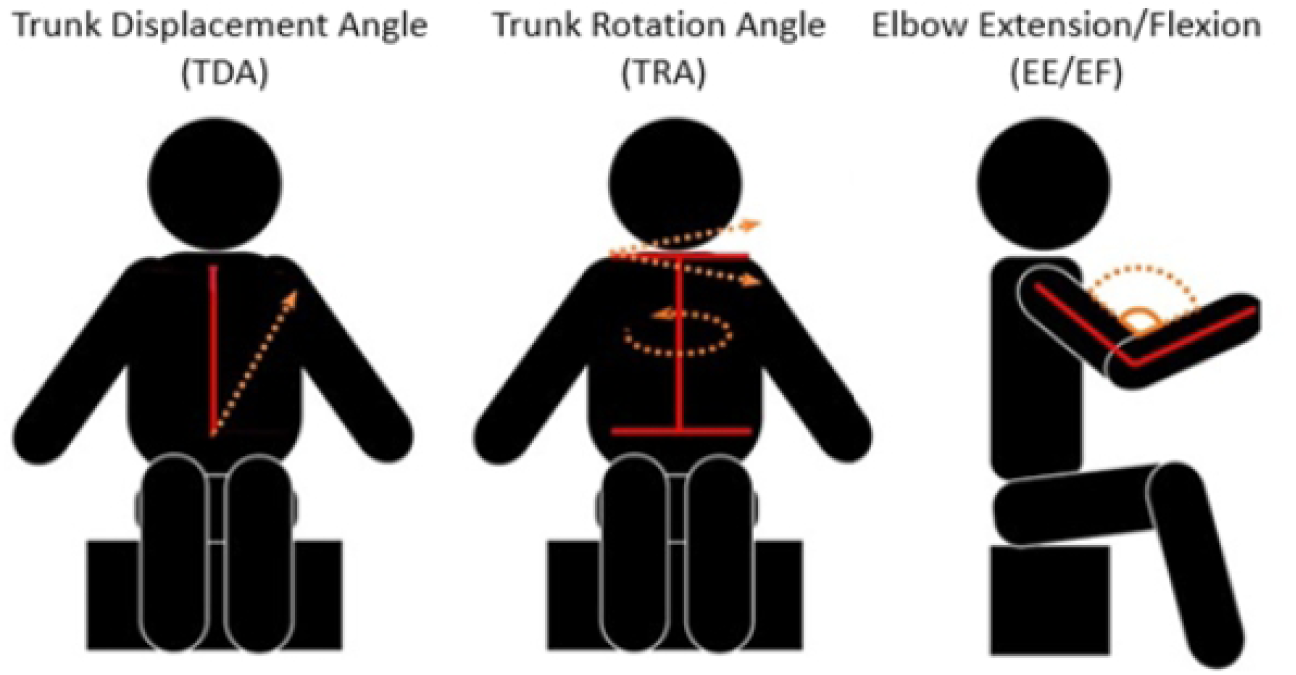
Graphical Depiction of Kinematic Variables. Note. The red lines represent the vectors, and the orange lines indicate possible movement directions. For TDA, the red line represents the median position

For the validation part we examined the elbow angle (Figure 5.3). This metric was also used for the secondary analyses where we examined movement time, velocity, and variability during reach, grasp, and transport phases. Reach phase elbow extension was analyzed as the joint angle change from rest to full extension, defined when the hand crosses a 100-degree threshold toward 95% maximum extension. Transport phase elbow flexion was similarly assessed in reverse (from full extension to flexion). Grasp time was the duration of object manipulation without elbow retraction. Movement time was calculated from elbow motion initiation to the end of each target phase. Velocity was the rate of elbow joint angular change during each phase. Movement variability was assessed pre- and post-therapy using the median absolute deviation (MAD), normalized to the session-specific median: Normalized MAD = Median (|*X*−Median|)/Median.

**Figure 5.3.**
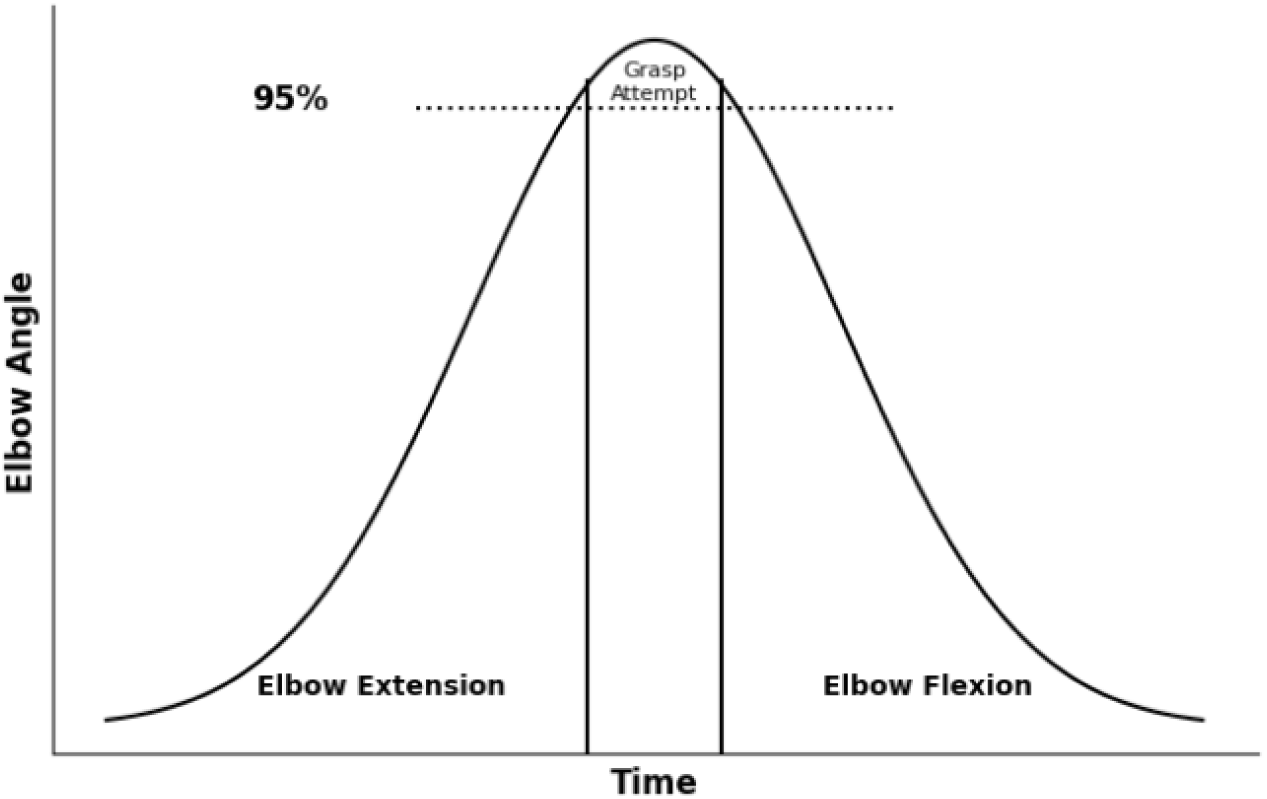
Illustration of Elbow Variable. Note. The y-axis represents elbow angle, and the x-axis represents time. A dashed line at 95% marks the grasp attempt.

For both 3DMA and video data, we used the Platform for Extracting and Analyzing Clinical Kinematics (PEACK) implemented in Python. The software allows analysis of various kinematic parameters, including joint angles. Time series data for selected joints (left/right shoulders, elbows, wrists, nose, and neck) were interpolated with cubic spline to fill missing values and filtered using a 3rd-order Butterworth FIR lowpass filter (cutoff frequency: 6 Hz). A non-linear median filter (window length: 0.5 seconds or 15 samples at 30 fps) was used on video data to remove sharp spikes. The PEACK pipeline exported an Excel CSV file containing all variables for 3DMA and 2D video data. For the trunk recruitment study, out of the 14 AHA play activities, certain tasks were included based on the specific variable that did involve the movement of interest (Table 5.1). Following task exclusion, the median values of the remaining tasks were computed for each participant, timepoint, and variable and exported to SPSS V21.

**Table 5.1.** AHA Tasks Included in Trunk Recruitment Study.

| **Task** | **TDA/TRA** | **EE/EF** |
| --- | --- | --- |
| Car in tube |  |  |
| Card sorting |  |  |
| Bottle with marbles inside | x |  |
| Putting on medal | x | x |
| Cooking using pan with lid | x |  |
| Crown/hat | x | x |
| Clapping with metal cymbals | x | x |
| Throwing paper | x |  |
| Drawing with markers | x |  |
| Opening music machine |  |  |
| Clapping with wood cymbals | x | x |
| Putting on bracelets | x | x |
| Opening music box |  |  |
| Manipulating pop up toy |  |  |

Note. x indicates inclusion of a task in variable calculation.

##### Temporal Synchronization for Validation Part

Because both data collection systems were manually initiated, temporal synchronization between the two data streams was initially imperfect. To correct this, we performed cross-correlations to identify the precise time lag between systems. This time lag was then computationally removed by appropriately shifting one data stream relative to the other.

#### Statistical Analysis

For the validation Bland-Altman analysis assessed agreement between OpenPose and the VICON system. Violations in normality, variance, and sphericity tests led to non-parametric analyses for trunk study. Between-subject differences were analyzed using Mann-Whitney U tests, and within-group differences with Wilcoxon signed-rank tests. Relationships were examined with Spearman ranked correlations. Effect sizes were calculated for all comparisons. Alpha level was set at p < 0.05.

## 5.3 Results

### 5.3.1 Validation Study

#### Agreement between 3DMA (VICON) and 2DMA (OpenPose)

Bland-Altman plots showed excellent agreement for TDA, with minimal bias (mean difference: –1.33°; −1.23°) and tight limits of agreement in both unimanual (–2.51° to −0.15°) and bimanual (–3.41° to 0.95°) tasks. TRA demonstrated moderate agreement, with a larger bias in the unimanual task (mean difference: – 4.19°, LoA: –6.33° to –2.05°) compared to bimanual (mean difference: –1.74°, LoA: –3.55° to 0.07°). Right elbow angles showed systematic bias with higher 3DMA values, especially in unimanual movements (mean difference: 56.59°, LoA: 21.92° to 91.25°) compared to bimanual (mean difference: 41.70°, LoA: 16.99° to 66.40°). Left elbow angles also favored 3DMA, but with smaller bias and reduced variability (unimanual mean difference: 17.34°, LoA: –11.92° to 46.60°; bimanual mean difference: 9.75°, LoA: –9.81° to 29.30°). Generally, higher 3DMA values reflect a tendency for the 2DMA system to underestimate joint angles compared to the 3DMA system (Figure 5.4).

**Figure 5.4.**
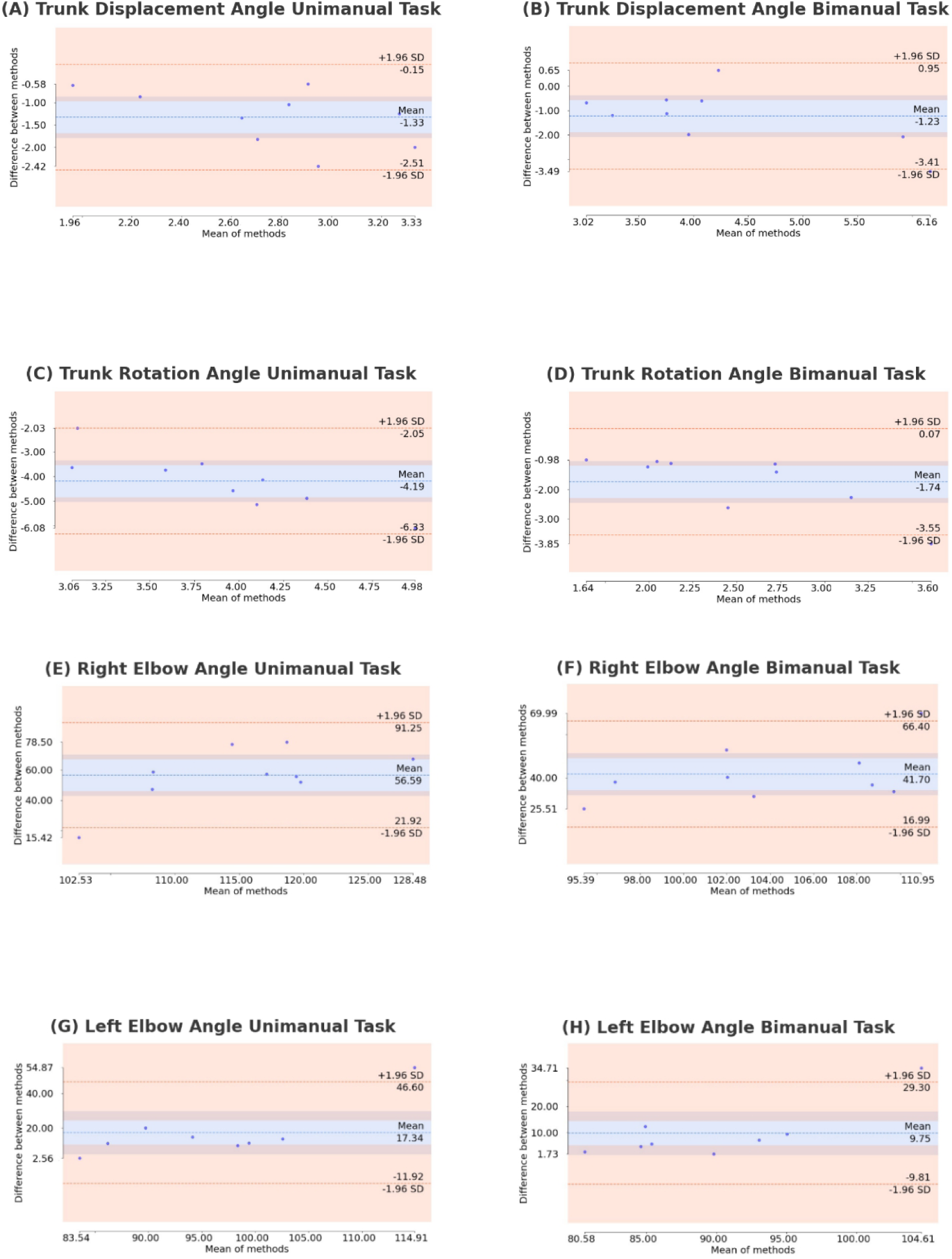
Agreement Between Methods in Unimanual and Bimanual Tasks. Note. The Bland-Altman plot provides a comparison of agreement between the OpenPose and VICON method, the mean difference and limits of agreement are expressed in degrees.

#### Relationship between 3D (VICON) and 2DMA (OpenPose)

TDA showed strong agreement, with slightly higher correlations in the bimanual task (r = 0.77; 0.80). TRA demonstrated moderate correlations (r = 0.64; 0.56). Right elbow angles showed weaker correlation unimanually (r = 0.45) and stronger correlation bimanually (r = 0.70). Left elbow angles showed consistently strong correlations, particularly in unimanual condition (r = 0.88; 0.74) (Table 5.2).

**Table 5.2.**
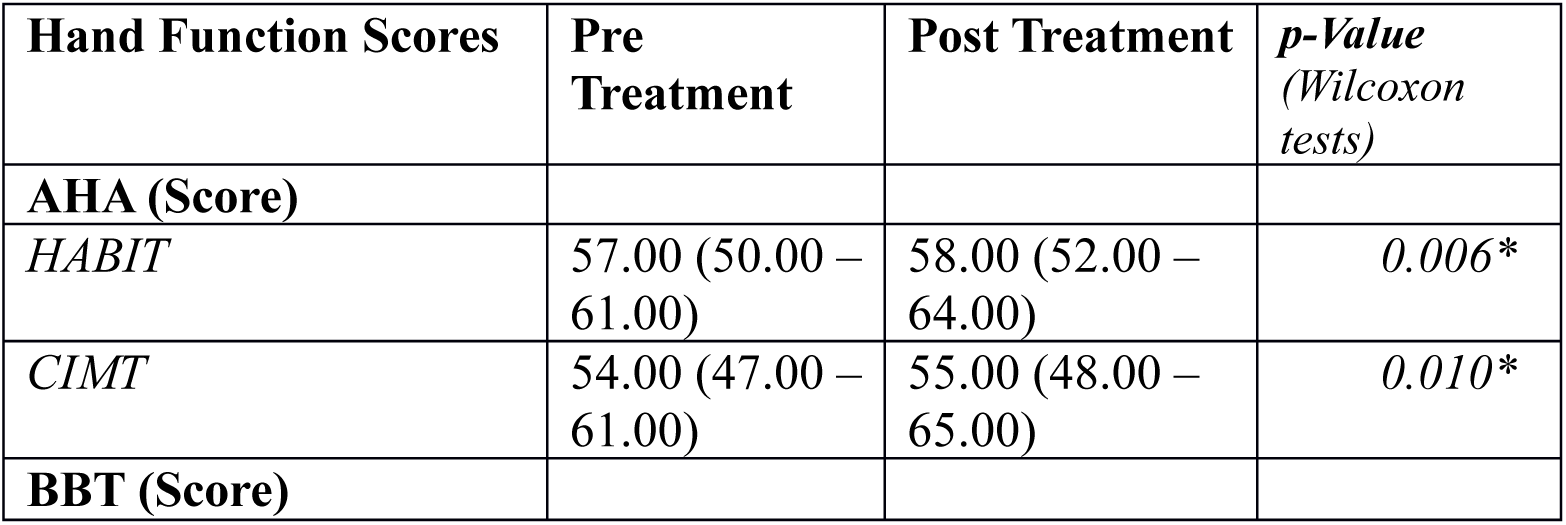
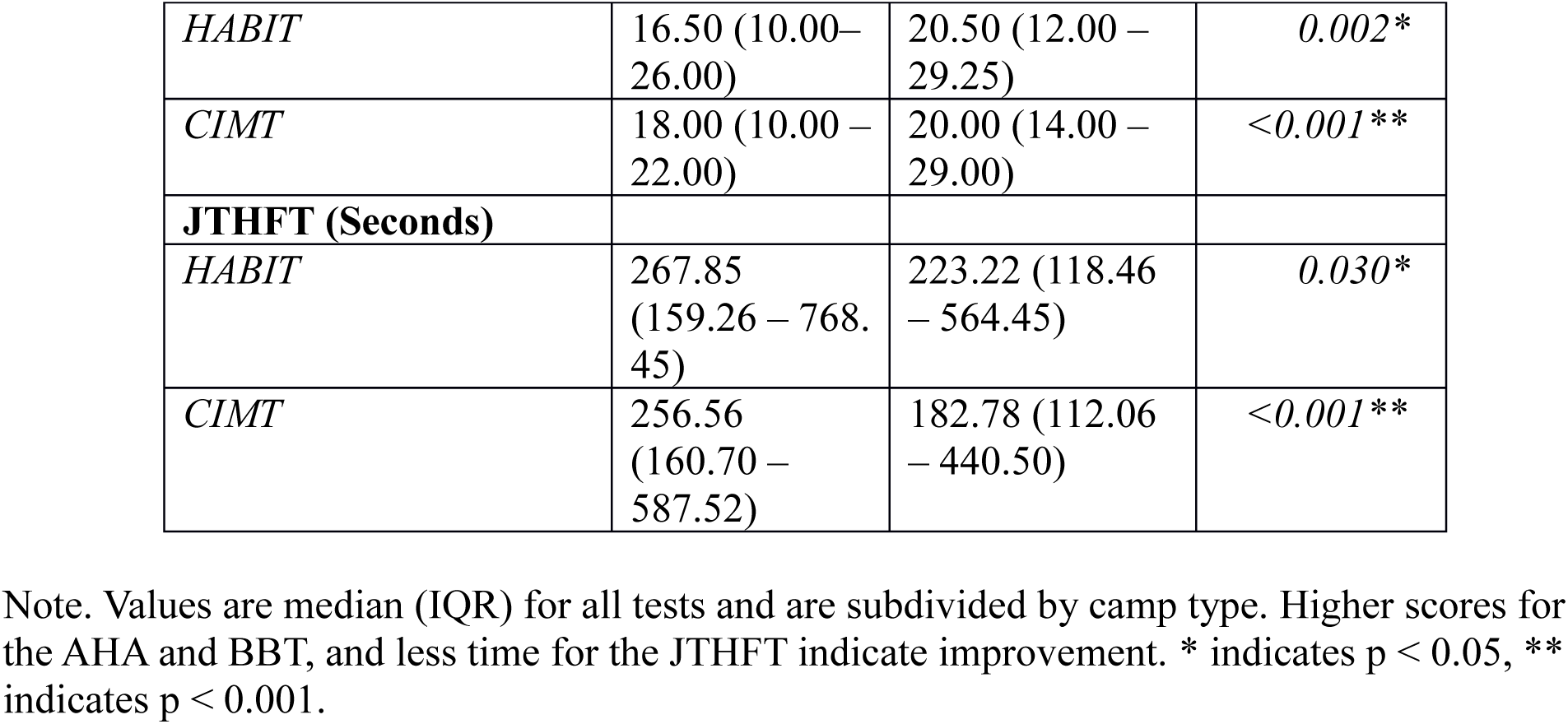
Hand Function Scores Pre and Post Treatment.

### 5.3.2 Trunk Recruitment Study

#### Baseline Analyses

*Missing Data.* Of the 50 children who participated in the camps (2015–2018), kinematic data could not be extracted for four, resulting in a final sample of 46 (22 CIMT, 24 HABIT). For the secondary analysis, data from 40 children (CIMT = 19; HABIT = 21) were analyzed after excluding data affected due to segmentation challenges (e.g., arm overlap and occlusion).

#### Hand Function Scores

Baseline AHA, BBT, and JTHFT scores were compared between CIMT and HABIT using the Mann-Whitney U test, revealing no significant pre-treatment differences. Wilcoxon signed-rank tests showed significant improvements in AHA, BBT, and JTHFT within both CIMT and HABIT groups. Mann-Whitney U tests on percent change scores showed no significant differences in improvement between CIMT and HABIT (see Table 5.2).

No significant baseline differences in TDA or TRA were observed between CIMT and HABIT treatments (Mann-Whitney U Tests) (Table 5.3).

**Table 5.3.** Trunk Variables Pre and Post Treatment.

| **Trunk Variables** | **Pre Treatment (degrees)** | **Post Treatment (degrees)** | **Percentage change** |
| --- | --- | --- | --- |
| **TDA** |  |  |  |
| *HABIT* | 8.59 (6.87 - 9.28) | 7.56 (6.70- 8.77) | -12.00 % |
| *CIMT* | 7.56 (7.22- 8.59) | 7.91 (6.87 - 9.45) | 4.54% |
| **TRA** |  |  |  |
| *HABIT* | 16.85 (13.92- 21.14) | 18.05 (13.06- 20.11) | 7.14 % |
| *CIMT* | 14.96 (11.00- 19.08) | 15.81 (12.72 - 18.56) | 5.75 % |

Note. Values are median (IQR) expressed in degrees for all tests and are subdivided by treatment type.

#### AIM 1. Exploring Pre-Treatment Relationships between Trunk Contribution and Hand Function

Spearman correlation revealed no significant relationships between trunk variables (TDA, TRA) and hand function measures (AHA, BBT, JTHFT) at baseline (TDA-AHA ρ = −0.08, p = 0.581; TDA-BBT ρ = .001, p = 0.99; TDA-JTHFT ρ = −0.028, p = 0.85; TRA-AHA ρ = −0.08, p =0.57; TRA-BBT ρ = 0.07, p = 0.66; TRA-JTHFT ρ = 0.02, p = 0.88); thus, regression analyses were not conducted. A strong correlation between TDA and TRA pre-scores (r = .635, p < 0.001) suggests these trunk movements occur in conjunction (Figure 5.5). However, Spearmen correlation with the ratio score of TDA/TRA and the AHA, BBT, and JTHFT baseline scores were not significant (AHA ρ = −0.03, *p= 0.86*; BBT: ρ = −0.05, *p = 0.75;* JTHFT: ρ = −0.03, *p = 0.84*).

**Figure 5.5.**
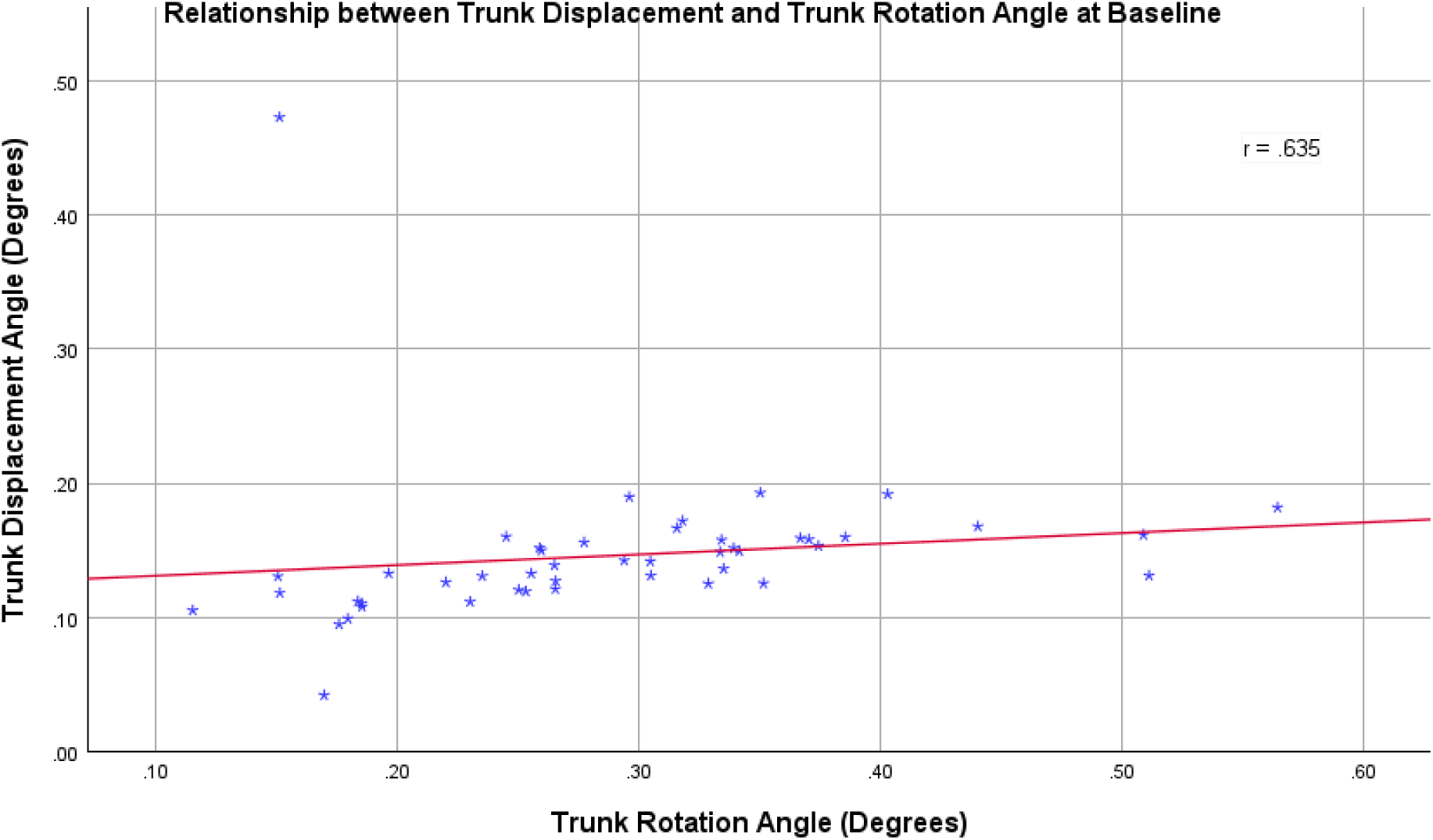
Relationship Between TDA and TRA at Baseline.

#### Secondary Analysis: Relationships Between Movement Metrics, Hand Function, and Trunk Variables at Baseline

*Prehension Variables at Baseline.* Mann-Whitney U tests showed no significant baseline differences between groups for any prehension variables (Figure 5.6).

**Figure 5.6.**
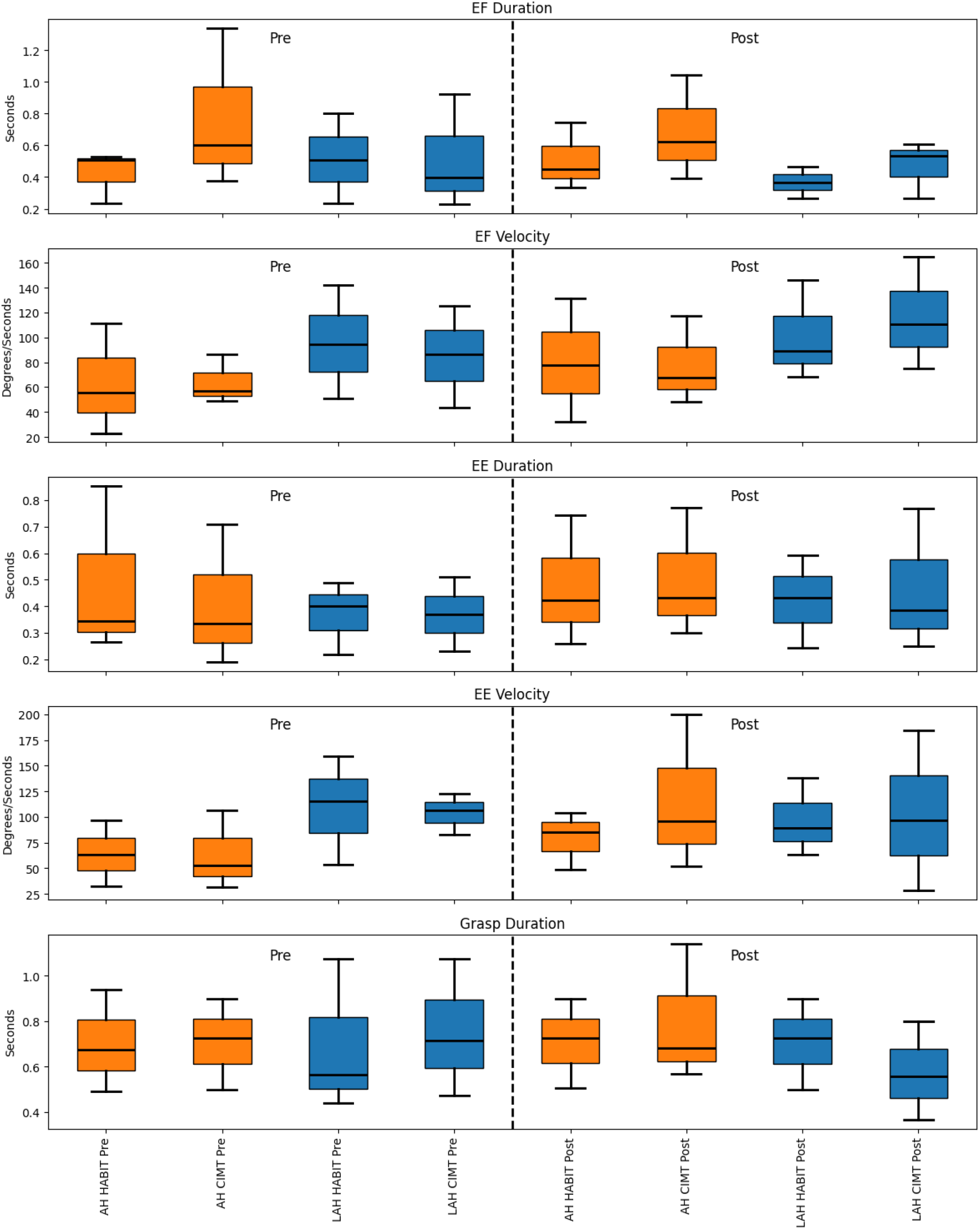
Comparison of Prehension Variables for Both Hands Pre and Post Treatment. Note. Boxplots display the distribution of prehension metrics, with the central line representing the median, the IQR showing the middle 50% of the data, and whiskers extending to 1.5 times the IQR. and Orange are more affected hand (AH) and blue are less affected hand (LAH) values.

#### Association Between Prehension, Hand Function, and Trunk Variables

Spearman correlations showed no significant relationships at baseline between hand function (AHA, BBT, JTHFT), trunk variables (TDA, TRA), and prehension metrics.

#### AIM 2: Changes in Trunk Contribution During AHA After Intensive Motor Learning

The Mann-Whitney U test on the change score of TDA between treatment groups (HABIT vs. CIMT) revealed a significant difference (p = .002), with a large effect size (r = 0.546). The median TDA in the HABIT group decreased from 8.59 to 7.56, while the CIMT group showed a small increase (pre-treatment median = 7.56, post-treatment median = 7.91) (Figure 5.7). The Mann-Whitney U test on TRA between treatment groups (HABIT vs. CIMT) revealed no significant difference *(p= 0.44).* Since there we no significant changes for TRA, further analyses with TDA and TRA change scores were not conducted.

**Figure 5.7.**
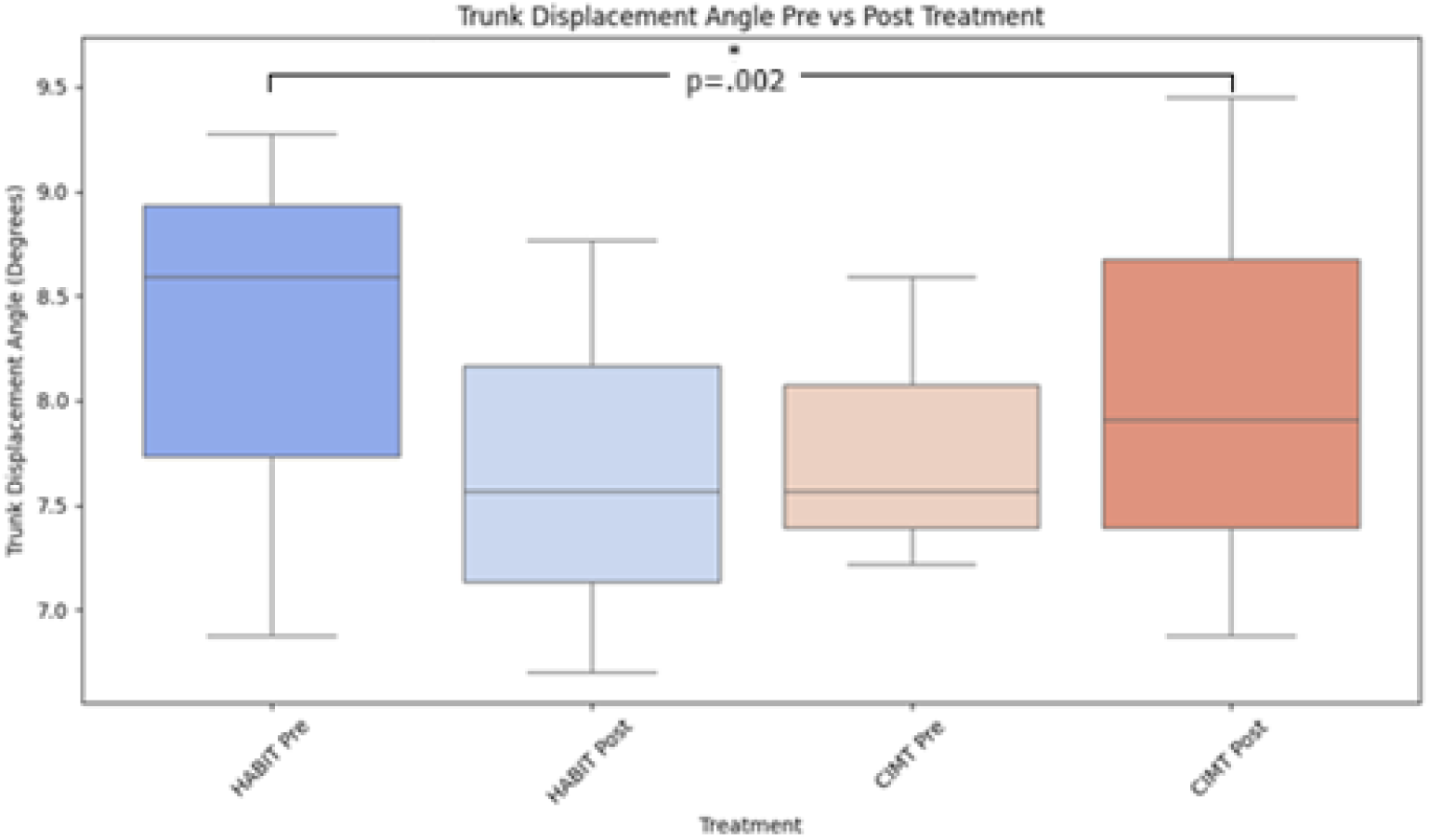
Boxplots Displaying TDA Pre and Post Treatment by Intervention Type. Note. The central line represents the median, the IQR showing the middle 50% of the data, and whiskers extending to 1.5 times the IQR. Significances are indicated by a * (p<.05).

#### Secondary Analysis: Impact of CIMT and HABIT on Movement Time, Velocity, and Variability of More AH

*Movement Time and Velocity.* Mann-Whitney U tests on change scores for reach, grasp, and transport phases showed no significant differences between treatment groups. Wilcoxon signed-rank tests within each group also revealed no significant changes across phases (see Table 5.4).

**Table 5.4.** Comparison of Velocity and Duration of the more AH.

| Prehension Variables | Pre Treatment Median (25th - 75th) | Post Treatment Median (25th - 75th) | *p value* (Mann Whitney Test) |
| --- | --- | --- | --- |
| Elbow Extension Angle Velocity (degree/sec) |  |  |  |
| *HABIT* | 67.97 (33.61- 105.59) | 77.75 (47.24 - 101.44) | *0.155* |
| *CIMT* | 52.86 (31.31 - 106.43) | 93.07 (52.92 - 190.78) |  |
| Elbow Flexion Angle Velocity (degree/sec) |  |  |  |
| *HABIT* | 61.41 (23.30 - 110.45) | 73.87 (32.65 - 129.64) | *0.320* |
| *CIMT* | 56.91 (48.61 - 86.53) | 82.30 (48.89 - 127.70) |  |
| Elbow Extension Duration (sec) |  |  |  |
| *HABIT* | 0.35 (0.27 - 0.85) | 0.45 (0.27 - 0.87) | *0.744* |
| *CIMT* | 0.33 (0.19 - 0.71) | 0.433 (0.30 - 0.74) |  |
| Elbow Flexion Duration (sec) |  |  |  |
| *HABIT* | 0.52 (0.28 - 0.97) | 0.45 (0.33 - 0.75) | *0.392* |
| *CIMT* | 0.60 (0.38 - 1.34) | 0.61 (0.40 - 1.00) |  |
| *HABIT* | 0.65 (0.47 - 0.88) | 0.72 (0.48 - 1.03) | *0.565* |
| *CIMT* | 0.68 (0.57 - 1.14) | 0.79 (0.41 - 1.23) |  |

Note. Comparison of the medians and IQR's and corresponding p-values for the Mann–Whitney U tests for velocity and duration for the more AH.

#### Variability

Mann-Whitney U tests also revealed no significant between-group differences across all phases. Wilcoxon signed-rank tests showed no significant pre-to-post changes in variability during reach, grasp, or transport phases for either CIMT or HABIT (Table 5.5).

**Table 5.5.** Comparison of Variability of the more AH.

| Prehension Variables | Pre Treatment Median (25th - 75th) | Post Treatment Median (25th - 75th) | *p value* (Mann Whitney Test) |
| --- | --- | --- | --- |
| Elbow Extension Angle Velocity (degree/sec) |  |  |  |
| HABIT | 36.59 (18.99 – 62.38) | 32.85 (10.88 – 68.99) | 0.454 |
| CIMT | 34.68 (25.29 – 115.82) | 56.53 (19.62 – 103.99) |  |
| Elbow Flexion Angle Velocity (degree/sec) |  |  |  |
| HABIT | 39.49 (19.60 – 52.22) | 39.48 (25.45 – 69.61) | 0.797 |
| CIMT | 14.65 (6.44 – 38.77) | 36.89 (18.28 – 60.15) |  |
| Elbow Extension Duration (sec) |  |  |  |
| HABIT | 0.22 (.04 - .52) | 0.25 (.150 - .450) | 0.560 |
| CIMT | 0.17 (.06 - .40) | 0.21 (.08 - .40) |  |
| Elbow Flexion Duration (sec) |  |  |  |
| HABIT | 0.26 (.08 - .42) | 0.20 (.12 - .47) | 0.416 |
| CIMT | 0.24 (.10 - .80) | 0.27 (.06 - .44) |  |
| Grasp Duration (sec) |  |  |  |
| HABIT | 0.20 (.06 - .43) | 0.27 (.08 - .48) | 0.968 |
| CIMT | 0.31 (.08- .63) | 0.35 (.09 - .72) |  |

Note. Comparison of the medians and IQR's and corresponding p-values for the Mann -Whitney U tests for variability of the more AH.

### 5.3.3 Exploratory Analysis of Elbow Extension Excursion as a Proxy for Trunk Compensation

Initial comparisons of trunk movement were conducted over the entire movement cycle, rather than isolating the prehension phase. It is possible that by including both the forward reach and return phases, the computation washed out meaningful findings, particularly since compensation is most likely to occur during the forward phase, when the child is actively reaching. To better isolate this phase and examine its relationship to hand function we computed elbow extension excursion as a surrogate marker for trunk involvement, using the formula: *Elbow Extension Excursion = Average Elbow Extension Velocity AH × Movement Duration AH* Spearman correlations revealed no significant relationships between elbow extension excursion and hand function measures (AHA, BBT, JTHFT) at baseline (Figure 5.8).

**Figure 5.8.**
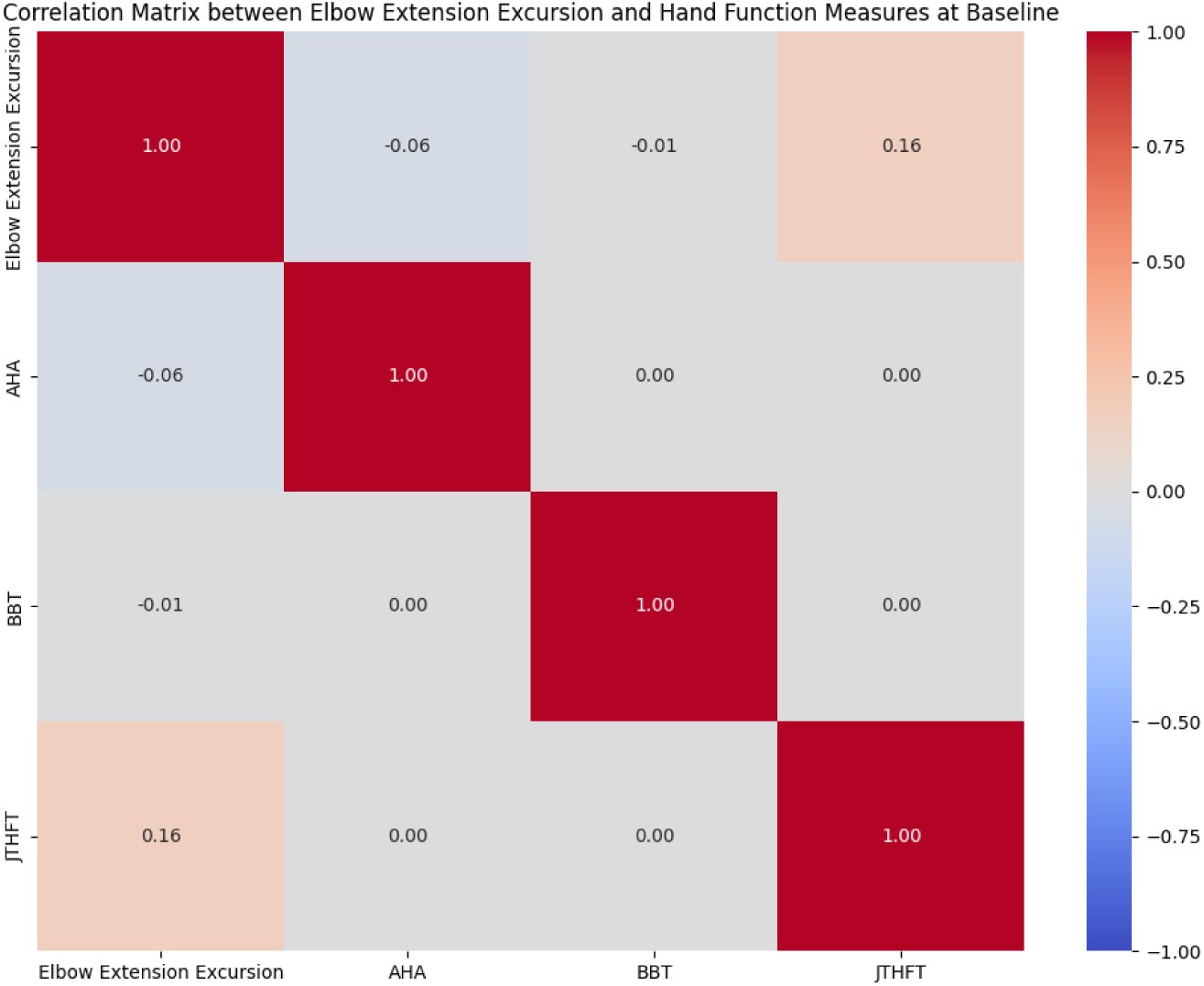
Correlation Matrix of Associations between Elbow Excursion and Hand Function Measures. Note. Matrix displays color-coded spearman correlation coefficients between elbow extension excursion and hand function measures. Diagonal values are self-correlations with values equal to 1.

The Mann-Whitney U test on the change score of elbow extension excursion between treatment groups (HABIT vs. CIMT) revealed no significant difference (Table 5.6). Therefore, no further analyses of the elbow extension excursion change scores were conducted.

**Table 5.6.**
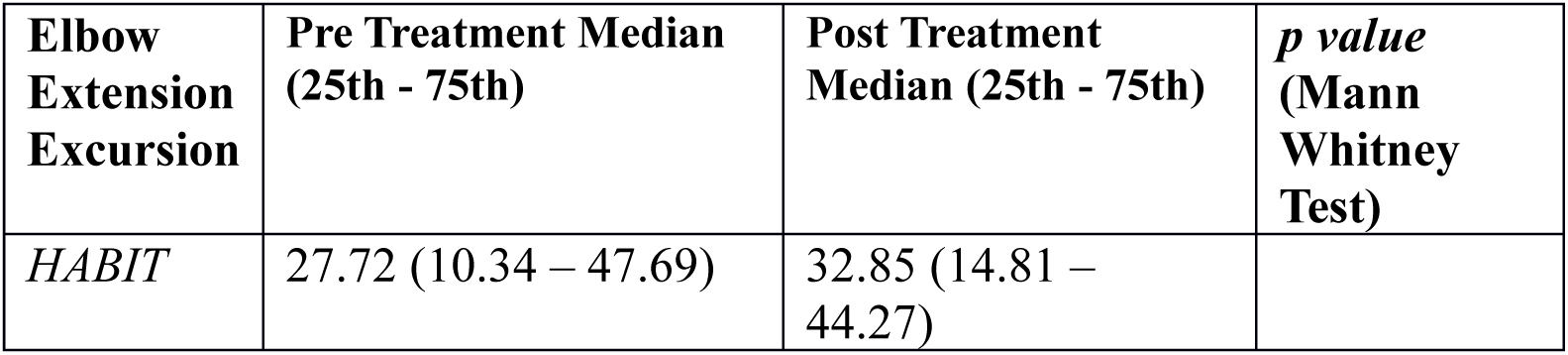
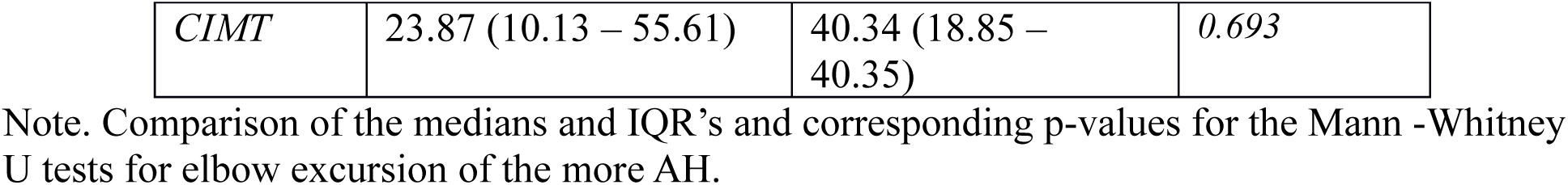
Comparison of Elbow Excursion of the more AH.

| Elbow Extension Excursion | Pre Treatment Median (25th - 75th) | Post Treatment Median (25th - 75th) | *p* value (Mann Whitney Test) |
| --- | --- | --- | --- |
| *HABIT* | 27.72 (10.34 – 47.69) | 32.85 (14.81 – 44.27) |  |
| *CIMT* | 23.87 (10.13 – 55.61) | 40.34 (18.85 – 40.35) | *0.693* |

Note. Comparison of the medians and IQR's and corresponding p-values for the Mann -Whitney U tests for elbow excursion of the more AH.

This indirect metric is intended to reflect the amount of reaching accomplished via elbow extension, assuming that greater excursion indicates less involvement of trunk movement during reach initiation. However, consistent with our original metrics, this derived measure also showed no meaningful correlation with hand function outcomes (Figure 5.8). Given the exploratory nature of this analysis and the indirectness of the measure, caution is warranted in interpreting these results.

## 5.4 Discussion

In this study, we conducted a retrospective analysis of hand function videos to investigate trunk involvement in children with USCP. We validated the use of OpenPose for our purpose by comparing it against a gold-standard 3D motion capture system. Our results suggested that OpenPose was reliable in capturing gross motor functions such as trunk movements, though caution is needed when interpreting more refined, complex motions. Using OpenPose, we examined how TDA and TRA relate to upper extremity function. We found that trunk involvement and kinematic variables of prehension were not strongly correlated to measures of hand function. We also evaluated how CIMT and HABIT influenced these measures of trunk and hand function. Our results suggest that while hand function improved following both CIMT and HABIT, the two interventions had different influences on trunk use, with HABIT significantly reducing it.

### 5.4.1 Reliability of OpenPose Compared to VICON

We evaluated the reliability of OpenPose for estimating upper limb and trunk kinematics in typically developed adults by comparing it to VICON 3DMA. Our findings showed that OpenPose performs well for proximal, sagittal-plane movements, with decreasing accuracy for multi-planar, axial movements and distal joints, consistent with prior studies.

Previous studies comparing OpenPose to gold-standard systems like VICON have reported strong agreement for multi-camera setups during lower extremity tasks such as walking, running, jumping (Needham et al., 2021), cycling (Pagnon et al., 2022), and knee drop tasks (Ino et al., 2024), particularly for large-amplitude, non-rotational movements. Studies on OpenPose using a single camera are scarce but findings are promising. For instance, Ota et al., (2020) reported near-perfect agreement with the VICON system for trunk, knee, and ankle angles during sagittal-plane squats using a single camera. Studies comparing OpenPose’s accuracy to 3DMA for upper limb kinematics are especially scarce and we could only identify one relevant study that has directly compared the two. In this study, Nakano et al., (2020) reported reduced accuracy for distal joints in a multi-camera setup during a throwing task, with smaller errors at the shoulder and larger errors at the elbow and wrist.

Our findings are consistent with these results. TDA, a sagittal movement, showed strong agreement across both unimanual and bimanual tasks. In contrast, TRA, an axial, multi-planar movement showed only moderate reliability, reflecting the known limitations of 2D pose estimation for capturing complex motion (Schurr et al., 2017; Straub & Powers, 2022). The greatest discrepancies occurred in elbow angle estimates. Right elbow measurements during unimanual tasks showed large disagreement with VICON. Bimanual tasks showed some improvement, and left elbow angles were more accurate overall but still fell short. However, the particularly low accuracy on the right side suggests that camera positioning and occlusion may have contributed to the error. Similarly, Nakano et al. (2020) found reduced accuracy for distal joints and attributed larger errors to frame-by-frame tracking failures, such as mislabeling objects or confusing limb segments. They suggested that incorporating an error-correction system could help OpenPose achieve acceptable accuracy for use in human movement science.

Although our findings align with previous work, the clinical relevance of our results remains uncertain, especially since there are no established minimum clinically important differences for trunk or reaching kinematics in USCP. Nonetheless, OpenPose shows promise as a scalable, low-cost tool for extracting gross motor kinematics from single videos. With further refinement, marker-less motion capture could help bridge the gap between standardized assessments and objective movement analysis in pediatric rehabilitation.

### 5.4.2 Absence of Pre-Treatment Associations Among Trunk, Prehension, and Hand Function

We found no significant relationships between trunk variables, prehension variables, and hand function before children with mild to moderate USCP participated in either a CIMT or a HABIT summer camp. This is in contrast with adult stroke literature, where trunk recruitment is typically correlated with impairment severity (Levin et al., 2002; Michaelsen et al., 2004; Wee et al., 2014). One explanation for this differential finding is that children’s motor systems are still maturing. According to Dynamic Systems Theory (DST), movement emerges from the continuous interaction between the nervous system, musculoskeletal system, and environment (Thelen, 1989). In children with USCP, it might be that early neuromuscular constraints limit their ability to explore a range of motor strategies, resulting in the premature stabilization of a trunk-based strategy as a dominant attractor state. In contrast, typically developing children explore multiple strategies before choosing efficient patterns (Hadders-Algra, 2000). Sveistrup et al., (2008) found that trunk, head, and arm coordination mature by age 2–3 for some tasks, and by 4–5 years resembles adult patterns, though even at 10–11, subtle differences persist, suggesting that trunk control continues to evolve.

In children with USCP, these developmental trajectories are altered, with children often relying more on trunk movement during reaching throughout development. This is supported by findings that individuals with hemiplegia don’t use their full movement capacity when given the choice because trunk recruitment is less effortful, suggesting that trunk involvement is not purely compensatory but a learned strategy (Michaelsen et al., 2004; Schneiberg et al., 2010; Wee et al., 2014). In children with USCP, early reliance on trunk-based movement pattern may therefore be a self-organizing solution as dominant attractor state.

Additionally, the level of impairment severity of our participants likely affected results. Our sample consisted of participants with mild to moderate USCP, which may explain the limited dependence observed between the trunk and hand function. Supporting this, Mailleux et al., (2017) found that milder impairments were associated with greater trunk stability. These results highlight the importance of tailoring interventions to the individual’s impairment level, as children with more severe motor deficits may rely more heavily on trunk compensation and could respond differently to treatments like CIMT or HABIT.

We observed a strong baseline correlation between TDA and TRA, suggesting that trunk movements are internally coordinated, even if it is not directly related to hand function. Trunk displacement and rotation often operate as a functional unit in both typically developing individuals and those with neuromotor impairments (Heide et al., 2003; Levin et al., 2002). Biomechanically, trunk displacement facilitates arm transport (Levin et al., 2002) while trunk rotation aligns the torso and shoulders toward the target (Cirstea & Levin, 2000). This coordination is especially evident in children with CP, who need to adjust their movement strategies due to their limited distal ability (Schneiberg et al., 2010). For example, in adults with post-stroke hemiplegia, the hand is often stabilized by proximal trunk and scapular movements, while elbow and hand joints remain relatively fixed (Cirstea & Levin, 2000; Jones, 2017). From a DST perspective, the observed correlation may reflect a stable attractor state that is a self-organized solution given the neuromotor constraints that helps achieving task goals in children with USCP.

Interestingly, we found no significant correlations between kinematic prehension measures and clinical hand function assessments. This is consistent with prior research (Gordon et al., 2011; Ratnadurai-Giridharan et al., 2024), which reported similarly weak or absent correlations. These findings highlight that clinical and kinematic measures may assess different dimensions of motor control: clinical assessments capture task success or performance, while kinematic analyses capture how the movement is performed. This underscores the limitations of relying on a single outcome measure to evaluate intervention effects. A child may demonstrate clinical improvement while adopting alternative movement strategies. Thus, multi-dimensional assessments that capture performance, capacity, and movement quality are essential to fully understanding motor function and for tailoring interventions effectively in children with USCP.

### 5.4.3 Intervention-Specific Trunk Adaptations Post Treatment

Both CIMT and HABIT improved hand function, but their effects on trunk recruitment were different. While HABIT led to a significant reduction in trunk displacement, CIMT did not. The decrease in HABIT aligns with its goal of promoting bilateral coordination and the functional use of both hands (Gordon et al., 2007). By integrating both hands more effectively, these children might have learned to redistribute motor demands across both upper limbs, transitioning to attractor state that reduces the need for proximal recruitment. Alternatively, the decrease in trunk displacement post-HABIT may be explained by the continued participation of less affected hand during more challenging parts of a task, leading to a more efficient task execution. In CIMT, the trend towards an increase in TDA likely reflects the intensive unilateral demand that it places on the more affected upper extremity (Charles et al., 2006). Forced to only use the more affected upper limb, children may recruit the trunk when distal control is insufficient to complete the task. While often viewed as maladaptive compensation, this can be seen through DST as an adaptive reorganization for maintaining stability under the constraints put on the body.

These findings partly align with existing literature. Our results for HABIT are consistent with Hung et al. (2018, 2020), who found reduced shoulder displacement following 90 hours of HABIT in children with USCP. In the first study, seven children with USCP received 90 hours of home-based HABIT. The study found that the children showed a decrease in trunk involvement as measured by a bimanual drawer opening task after the intervention (Hung et al., 2018). In the later study, 10 children with USCP participated in a camp where they received 90 hours of HABIT. The study found that after the intervention, the children showed a decrease in trunk involvement as measured by a unimanual reach-grasp eat task (Hung et al., 2020). However, our CIMT findings contrast with Hung et al. (2020), who found that 90 hours of CIMT reduced shoulder displacement in 10 pediatric individuals with USCP. In that study, children performed a reach-grasp-eat task while being recorded with a 3D motion analysis system. Together, these findings show the importance of individualized intervention design.

As Cirstea and Levin (2000) proposed, patients who are falling below a specific threshold in elbow extension, shoulder flexion, and adduction might benefit to compensating for the loss of ability by utilizing their trunk. In contrast, patients above this threshold, despite their tendency to use trunk compensatory strategies, may still retain the ability to use the ranges of motion in their arm joints effectively and benefit from appropriate therapy to enhance and recover lost ranges of joint motion. This framework is supported by the findings of Sakzewski et al., (2011) showing that those children with USCP with worse hand function experienced greater gains in unimanual capacity after motor learning-based interventions, suggesting that baseline motor capacity should be considered when selecting intervention approaches. They also highlight the need for multi-dimensional assessments that capture both performance outcomes and the quality of movement to evaluate intervention effects.

The lack of significant changes in TRA between pre- and post-intervention in both intervention groups could be due to two factors: TRA might be harder to change because it is a rotational movement that is multi-dimensional and requires coordination across several joints and muscles (Fujii et al., 2007), which might be more difficult to modify. Additionally, both of our interventions focused on the upper limb, especially the hand, which might have led participants to prioritize motor control improvements for arm and hand function, rather than focusing on refining trunk movement strategies. This interpretation is consistent with prior findings indicating that trunk displacement and rotation serve distinct roles in postural control and movement compensation (Yang et al., 2021).

### 5.4.4 Lack of Differential Prehension Changes After CIMT and HABIT

In our secondary analysis, we hypothesized that CIMT would reduce grasp duration more than HABIT, given its intensive focus on repetitive unimanual practice (Gordon et al., 2005). However, both groups showed only modest, non-significant reductions. This suggests that while coordination may improve with intervention, execution speed may not, possibly due to persistent challenges in motor planning or sensorimotor integration. This finding aligns with Hung et al., (2017), who observed improvements in motor control but not in movement time. Another explanation is the use of the speed–accuracy trade-off to prioritize accuracy over speed for task success (Fernani et al., 2017).

Although both groups exhibited increases in elbow velocity, these changes were not statistically significant, with the CIMT group showing an 80.8% increase and the HABIT group a 35.8% increase. This trend suggests that CIMT may facilitate movement speed through high-repetition, task-specific use of the more affected limb. In contrast, HABIT’s bimanual focus might promote improvements in coordination and bilateral integration without directly targeting velocity. However, given the lack of statistical significance, further studies with larger sample sizes are necessary to determine the efficacy of these therapies in enhancing movement speed.

Interestingly, movement time remained unchanged in both groups. This might be explained by the nature of the task in which the prehension variables were examined. The AHA is a performance test rather than a capacity test and conducted at a self-selected pace (Krumlinde-Sundholm et al., 2007). In DST terms, if the task constraints do not demand faster execution, the motor system may stabilize around solutions that optimize success without the pressure to accelerate (Newell & Irwin, 2021). Improvements in control and precision may therefore not immediately translate into shorter movement durations.

We also hypothesized that CIMT would reduce movement variability more than HABIT. However, neither group demonstrated significant reductions. While traditionally viewed as undesirable, variability is increasingly recognized as a functional component of motor learning that enables exploration and adaptation, facilitating the development of flexible motor strategies (Dhawale et al., 2017; López-Fernández et al., 2025). Visual inspection revealed that variability was not static in our sample but showed rather wide interquartile ranges. From a DST perspective, such variability would reflect a motor system that is still exploring and has not yet found a stable coordination pattern (Thelen, 1989). This interpretation of our results aligns with Hung et al. (2017), who found that spatial variability may decrease with training, while velocity variability can persist. These findings suggest that movement variability may require longer or more targeted intervention to stabilize, particularly in populations where compensatory strategies are deeply ingrained or underdeveloped.

### 5.4.5 Study Limitations and Future Directions

Our study primarily included children with mild to moderate impairments (MACS I–III), which may limit the generalizability of our findings. Children with more severe impairments (MACS IV–V) may show stronger or different trunk patterns, affecting the relationship between trunk movements and hand function. Future studies should include a wider range of impairment levels to assess whether differential effects can be found across the USCP spectrum.

Another factor to consider is the duration and intensity of the interventions. Short-term interventions may not provide sufficient time for participants to develop and consolidate new motor patterns, particularly for complex movements like trunk rotation. Extending the intervention period or increasing its intensity might help seeing more pronounced effects (Hsu et al., 2019).

Additionally, measurement constraints need to be considered. Our validation showed that OpenPose effectively tracked TDA with strong agreement and temporal correspondence with the gold-standard 3DMA system (VICON). Bland-Altman and time-series analyses showed minimal bias and high correlation. Prior research supports 2D motion capture as being comparable to 3D for sagittal movements (Scataglini et al., 2024), but the complexity of trunk-arm coordination in USCP may require more sensitive, task-specific tools. These technical issues highlight the need for a multi-camera system or standardized setup to capture complex, multi-plane motion accurately. Moreover, our assessments were conducted in a controlled lab setting, which may not fully reflect how children perform tasks in real life. While controlled conditions support consistency, real-world tasks involve greater environmental variability. Future studies should examine how intervention-driven movement adaptations translate to everyday settings.

## 5.5 Conclusion

This study provides new insights into the complex relationship between trunk movements, upper extremity function, and prehension in children with USCP, highlighting the distinct effects of CIMT and HABIT interventions. HABIT reduced trunk involvement, whereas CIMT tended to increase trunk use, highlighting the importance of individualized intervention selection based on each child’s motor abilities. The lack of direct correlations between trunk and hand function suggests that pediatric USCP does not follow predictable coordination patterns seen in adults post-stroke, reflecting the adaptability of the developing nervous system. Additionally, the validation of OpenPose as an alternative to traditional motion analysis systems is a methodological advancement, however refinements are needed for certain variables that are multiplanar. Most importantly, our findings contribute to informed decision-making in intervention allocation. Rather than eliminating all trunk recruitment, interventions should support self-organization by optimizing function within each child’s unique constraints, enabling more personalized and effective rehabilitation allocation.

## Data Availability

All data produced in the present study are available upon reasonable request to the authors

## Acknowledgements

We would like to thank all the families participating in our camps over the years as well as all interventionists for their hard work.

This study was funded by The Order of Malta.

## Notes

### Competing Interest Statement

The authors have declared no competing interest.

### Funding Statement

The study was funded by The Order of Malta

### Author Declarations

Ethics committee/IRB of Teachers College, Columbia University gave full ethical approval for this work

